# Analysis of early renal injury in COVID-19 and diagnostic value of multi-index combined detection

**DOI:** 10.1101/2020.03.07.20032599

**Authors:** Xu-wei Hong, Ze-pai Chi, Guo-yuan Liu, Hong Huang, Shun-qi Guo, Jing-ru Fan, Xian-wei Lin, Liao-zhun Qu, Rui-lie Chen, Ling-jie Wu, Liang-yu Wang, Qi-chuan Zhang, Su-wu Wu, Ze-qun Pan, Hao Lin, Yu-hua Zhou, Yong-hai Zhang

**Author notes:** These authors contributed equally to this work (Xu-wei Hong, Ze-pai Chi, Guo-yuan Liu). **Corresponding author:** Yong-hai Zhang; E-mial; Department of Urology, Shantou Central Hospital, Shantou 515031, Guangdong Province, People’s Republic of China.

## Abstract

**Objectives:** The aim of the study was to analyze the incidence of COVID-19 with early renal injury, and to explore the value of multi-index combined detection in diagnosis of early renal injury in COVID-19.

**Design:** The study was an observational, descriptive study.

**Setting:** This study was carried out in a tertiary hospital in Guangdong, China.

**Participants:** 12 patients diagnosed with COVID-19 from January 20, 2020 to February 20, 2020.

**Primary and secondary outcome measures:** The primary outcome was to evaluate the incidence of early renal injury in COVID-19. In this study, the estimated glomerular filtration rate (eGFR), endogenous creatinine clearance (Ccr) and urine microalbumin / urinary creatinine ratio (UACR) were calculated to assess the incidence of early renal injury. Secondary outcomes were the diagnostic value of urine microalbumin (UMA), α1-microglobulin (A1M), urine immunoglobulin-G (IGU), urine transferring (TRU) alone and in combination in diagnosis of COVID-19 with early renal injury.

**Results:** While all patients had no significant abnormalities in serum creatinine (Scr) and blood urea nitrogen (BUN), the abnormal rates of eGFR, Ccr, and UACR were 66.7%, 41.7%, and 41.7%, respectively. Urinary microprotein detection indicated that the area under curve (AUC) of multi-index combined to diagnose early renal injury in COVID-19 was 0.875, which was higher than UMA (0,813), A1M (0.813), IGU (0.750) and TRU (0.750) alone. Spearman analysis showed that the degree of early renal injury was significantly related to C-reactive protein (CRP) and neutrophil ratio (NER), suggesting that the more severe the infection, the more obvious the early renal injury. Hypokalemia and hyponatremia were common in patients with COVID-19, and there was a correlation with the degree of renal injury.

**Conclusions:** Early renal injury was common in patients with COVID-19. Combined detection of UMA, A1M, IGU, and TRU was helpful for the diagnosis of early renal injury in COVID-19.

## Introduction

Since December 2019, unexplained clustered pneumonia cases have started to appear in Wuhan, Hubei Province, China. It is identified as a pneumonia caused by a novel coronavirus infection.[1,2] The World Health Organization (WHO) named the coronavirus 2019-nCoV, which was the third coronavirus that infects humans from wild hosts across germlines after severe acute respiratory syndrome coronavirus (SARS-CoV) and Middle East respiratory syndrome coronavirus (MERS-CoV) and can cause severe pneumonia. It is also the seventh species of pathogenic human respiratory coronaviruses.[3,4] On February 11, 2020, the WHO named the novel coronavirus-infected disease2019 novel coronavirus disease shorted for COVID-19. With the spread of the epidemic, other cases in China and abroad have also occurred.[5] As of February 22, 2020, a total of 76,396 cases were diagnosed in China, including 11,477 severe cases and 2,348 deaths.

The main symptoms of COVID-19 are fever, dry cough, and fatigue. A few patients have symptoms of nasal congestion, runny nose, sore throat, and diarrhea. Severe patients usually develop dyspnea and / or hypoxemia one week after the onset of the disease. In severe cases, they quickly develop into acute respiratory distress syndrome (ARDS), septic shock, and multiple organ injury such as cardiac, hepatic, and renal injury.[6] The 2019-nCoV human infection cases combine the characteristics of the previous 6 coronaviruses, which can cause mild cases, easily lose vigilance, and can also cause severe cases, with a high mortality rate. [7] Multiple organ failure is the main cause of death of COVID-19. The latest research reports that the incidence of COVID-19 with organ dysfunction is about 33%, of which the acute renal injury is about 3 ∼ 7%. [8,9] Impaired renal function can lead to obstruction of excretion of metabolites and toxins in the body, which will adversely affect the maintenance of the electrolyte and acid-base balance of the human body. In addition, when renal function is severely damaged, uremia will occur and endanger life. Early detection of evidence of renal injury and timely effective interventions are of great significance for reducing complications and improving prognosis.

This study intends to use a number of laboratory test indexes, including serum creatinine, blood urea nitrogen, urine creatinine, urine microalbumin and urine microglobulin et al to comprehensively assess renal function and determine the incidence of COVID-19 with early renal injury. The related risk factors of COVID-19 with early renal injury were also analyzed.

## Materials and methods

### Patients

From January 20, 2020 to February 20, 2020, 12 patients with pneumonia of unknown cause were admitted to a tertiary hospital in Guangdong, China. Nasopharyngeal swab samples of all patients were tested positive for 2019-nCoV virus nucleic acid and confirmed to be infected with 2019-nCoV by Guangdong Province CDC (Center for Disease Control and Prevention). The disease diagnostic criteria and case classification were refer to the guidelines for diagnosis and treatment of COVID-19 (Trial Version 5) issued by China National Health Commission.[6]

### Data collection

A standardized case collection form was designed to collect laboratory data of the included patients. It mainly included the following items: (1) blood routine; (2) biochemical indexes of kidney, liver, and heart function; (3) coagulation function; (5) infection indexes.

### Evaluation of renal function

Serum creatinine (Scr), blood urea nitrogen (BUN), urine creatinine (Ucr), urine protein (PRO), urine microalbumin (UMA), α1-microglobulin (A1M), urine immunoglobulin G (IGU), and urine transferrin (TRU) were detected. The estimated glomerular filtration rate (eGFR), endogenous creatinine clearance (Ccr), and urine microalbumin/creatinine ratio (UACR) were calculated.

Among them, the eGFR was calculated according to the simplified MDRD formula modified by the Chinese.

eGFR=186×Scr(umol/L)^−1.154^×Age(year-old)^−0.203^(female×0.742)

(the Counahan-Barrat method was applied when the patient is younger than 14-year-old)

The Ccr was calculated according to Cockcroft’s formula.

Ccr=[140-Age(year-old)] ×weigh(t kg)÷[0.818×Scr(umol/L)](female×0.85)

In this study, two or more abnormalities of eGFR, Ccr and UACR were defined as early renal injury.

### Statistical methods

The area under curves (AUC) of receiver operating characteristic (ROC) were calculated for predictive analysis. Spearman rank correlation coefficient was used to analyze the linear correlation between two sets of continuous variables. It was considered statistically significant when the P value was less than 0.05. All statistical analyses were processed using SPSS 25.0 statistical software.

### Patient and public involvement

Patients were not directly involved in the design, planning and conducting of this study.

## Results

### Laboratory characteristics of the COVID-19 patients

Of the 12 patients with COVID-19, 2 were severe patients, 8 were general patients, and 2 were light patients. Laboratory testing items included blood routine, electrolyte, metabolism, heart, liver, kidney function indicators, coagulation function and infection indicators. As shown in Table 1, the common abnormal indicators (abnormal rate ≥50%) included: increased neutrophil ratio (50%), increased monocyte ratio (75%), hypokalemia (50%), hyponatremia (50%), hypoproteinemia (75%), and increased C-reactive protein (83.3%).

**Table 1.**
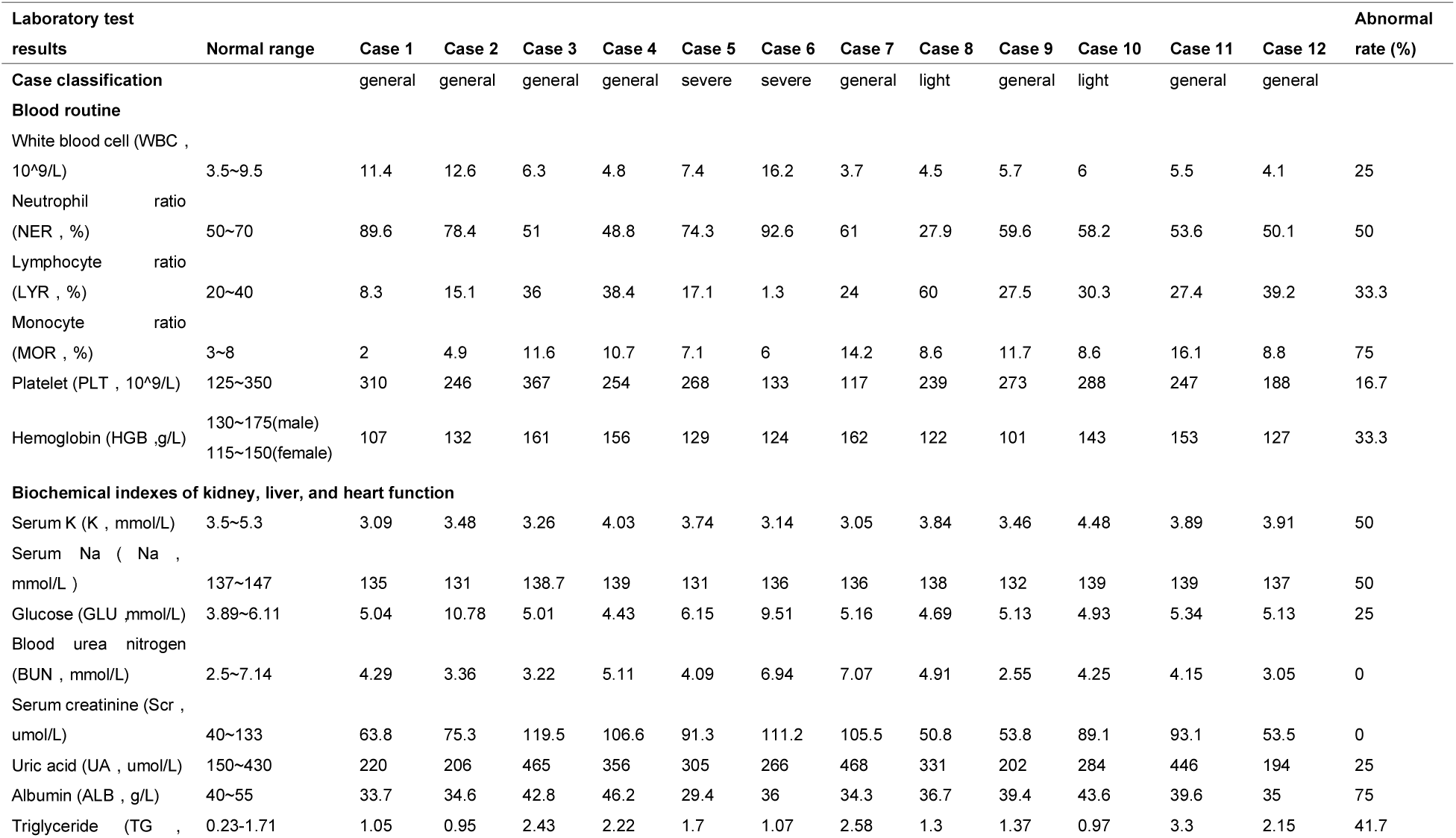
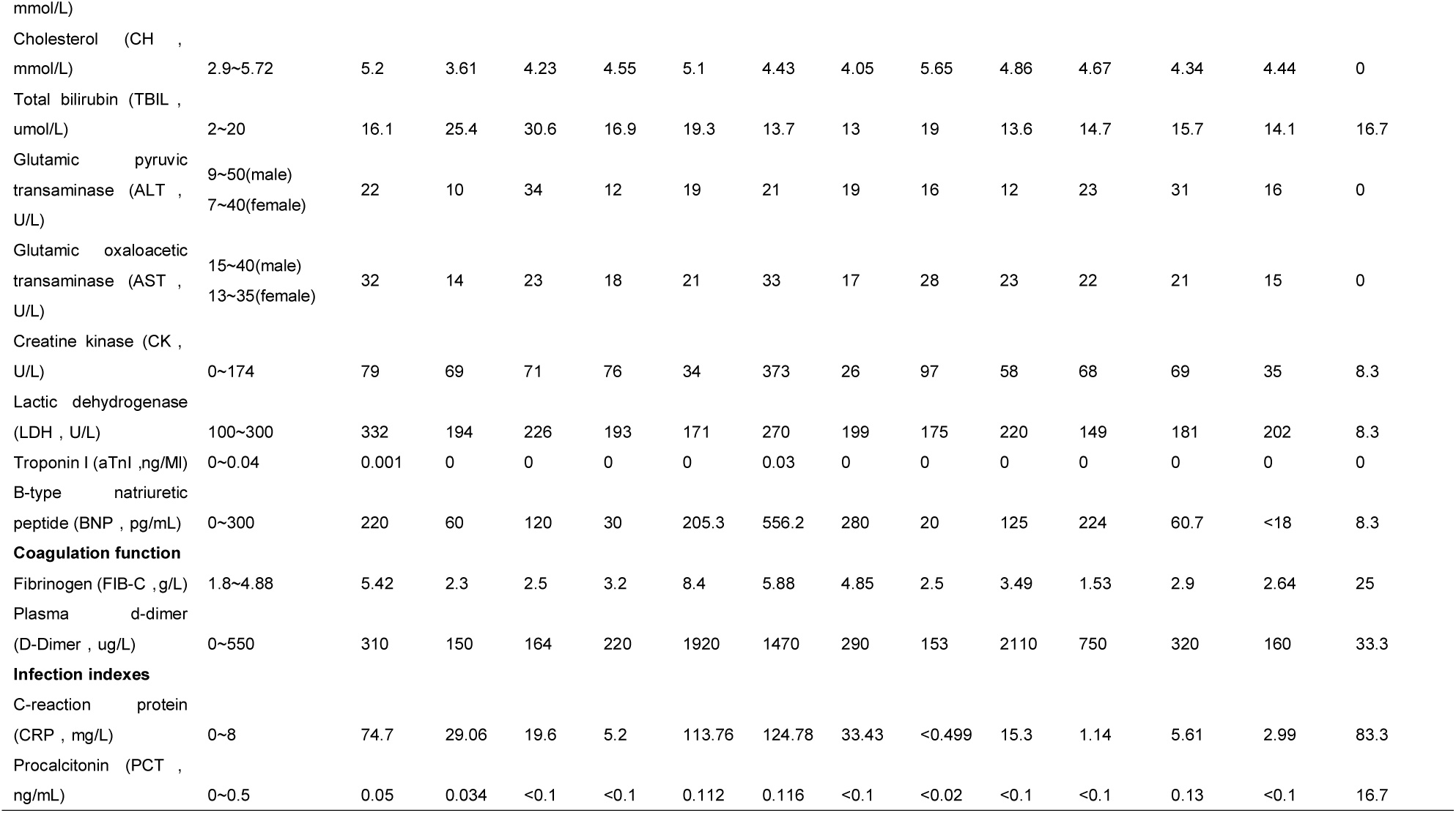
Laboratory test results of the COVID-19 patients

### Incidence of COVID-19 with early renal injury

As shown in Table 2, Scr and BUN were not significantly abnormal in 12 patients. However, the calculation of eGFR and Ccr showed that early renal injury was common in COVID-19 patients. Among them, 66.7% patients had reduced eGFR and 41.7% patients had reduced Ccr. Urinary microprotein test also confirmed that COVID-19 with early renal injury was common, with UACR> 30mg / g accounting for 41.7%. Among the 12 patients, the abnormally elevated rates of UMA, A1M, IGU, and TRU were 33.3%, 33.3%, 41.7%, and 16.7%, respectively. The positive rate of the four-index combined was 58.3%. The results suggested that case 3, 5, 6, and 7 were significant early kidney injury cases.

**Table 2.**
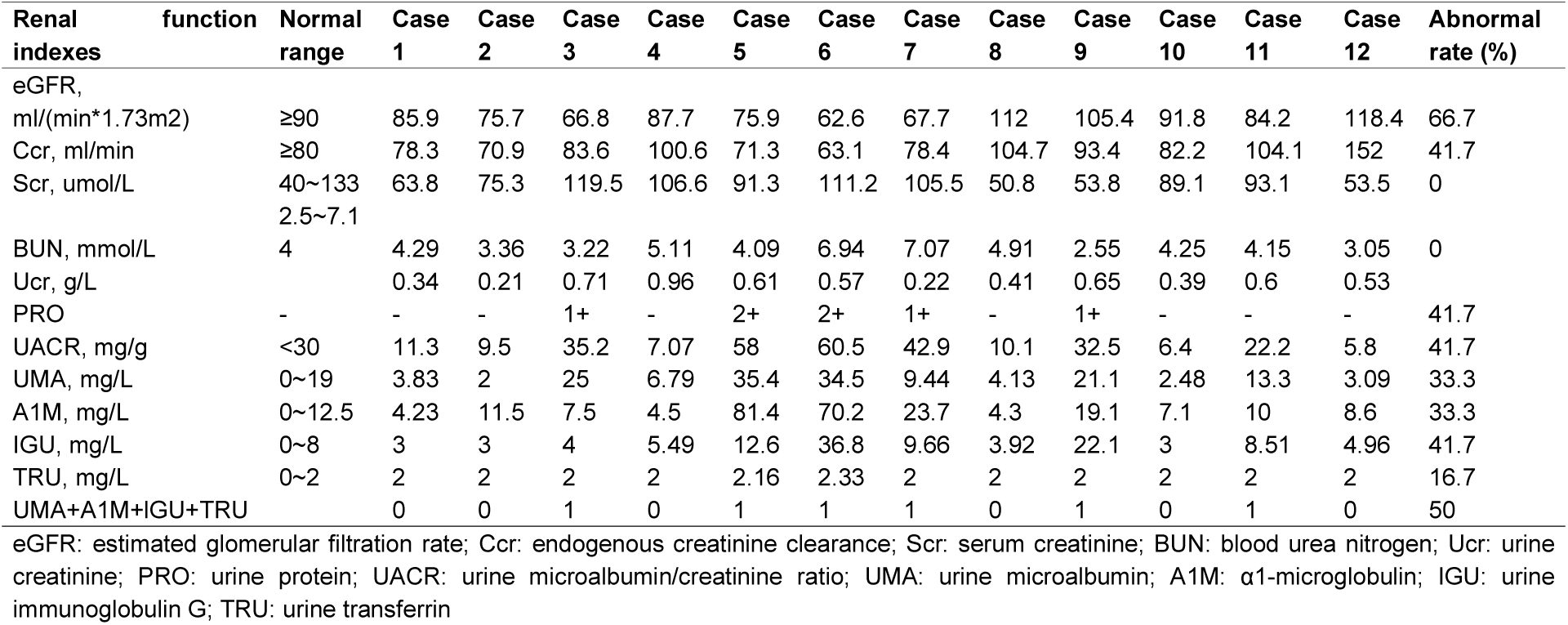
Renal function indexes of the COVID-19 patients

### Urinary microprotein detection in diagnosis of early renal injury in COVID-19

The AUC of UMA, A1M, IGU, and TRU for the diagnosis of early renal injury in COVID-19 were 0.813, 0.813, 0.750, and 0.750 respectively, while the AUC of the combined index of UMA + A1M + IGU + TRU was 0.875, suggesting that the multi-index combined was helpful for the diagnosis of early renal injury in COVID-19, as shown in Fig 1.

**Figure 1.**
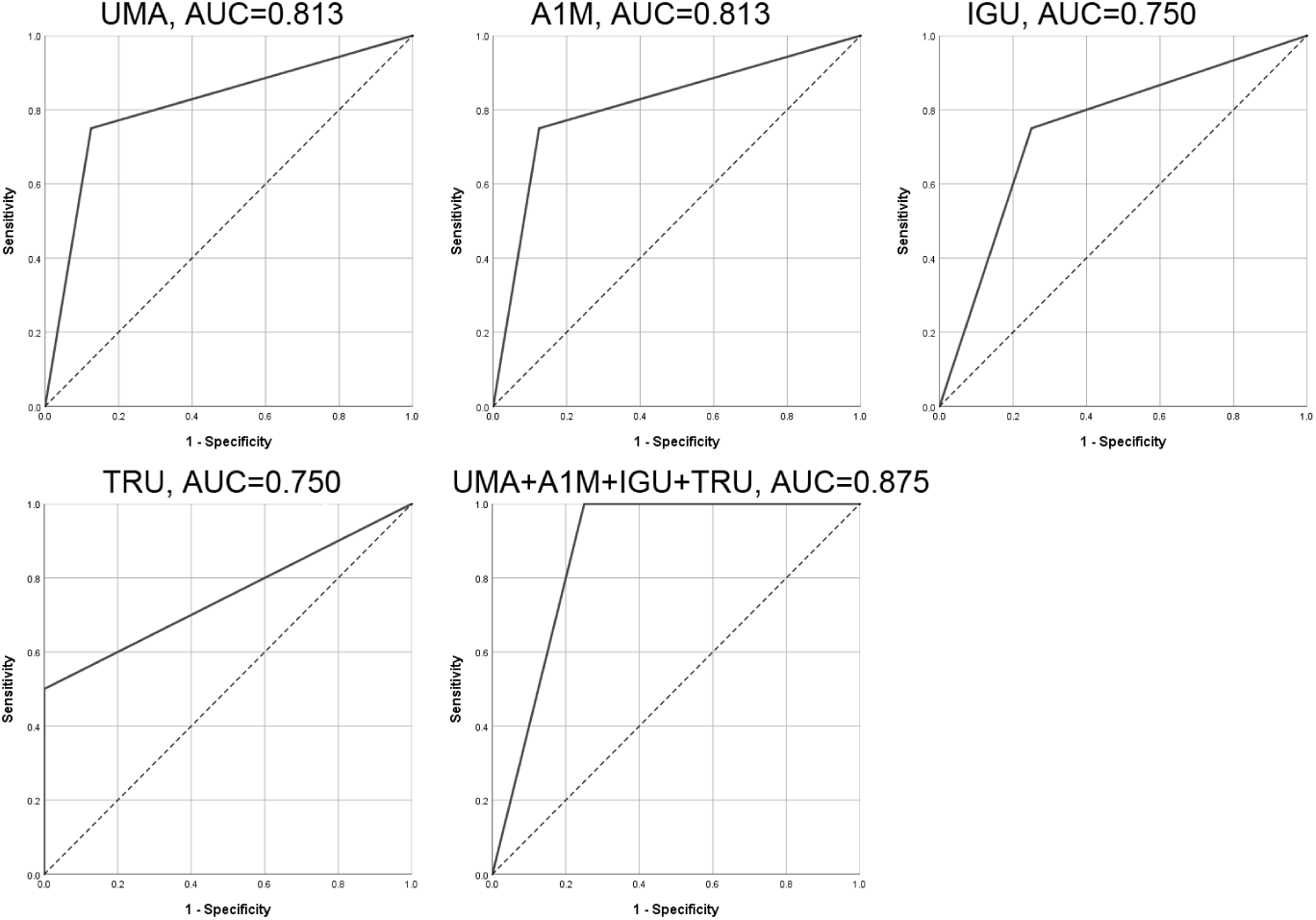
ROC curves of urinary microprotein (UMA, A1M, IGU, TRU and multi-index combined) in predicting early renal injury in COVID-19. (ROC: receiver operating characteristic; AUC: area under curve; UMA: urine microalbumin; A1M: α1-microglobulin; IGU: urine immunoglobulin G; TRU: urine transferrin)

### Factors related to early renal injury in COVID-19

In this study, Spearman correlation coefficients were used to evaluate the correlation between renal function indicators (eGFR, Ccr, and UACR) and the obviously abnormal biochemical indicators (C-reactive protein (CRP), neutrophil ratio (NER), monocyte ratio (MOR), serum potassium (K), serum sodium (Na), and albumin (ALB)). As shown in Table 3, it was found that eGFR was negatively correlated with CRP and NER, and positively correlated with serum K, Ccr was negatively correlated with CRP and NER, and positively correlated with MOR and serum Na, and UACR was positively correlated with CRP, and negatively correlated with serum K (Fig 2). The results suggested that the more severe the infection, the more obvious the early renal injury, and the early renal injury in COVID-19 can often cause hypokalemia and hyponatremia.

**Table 3.**
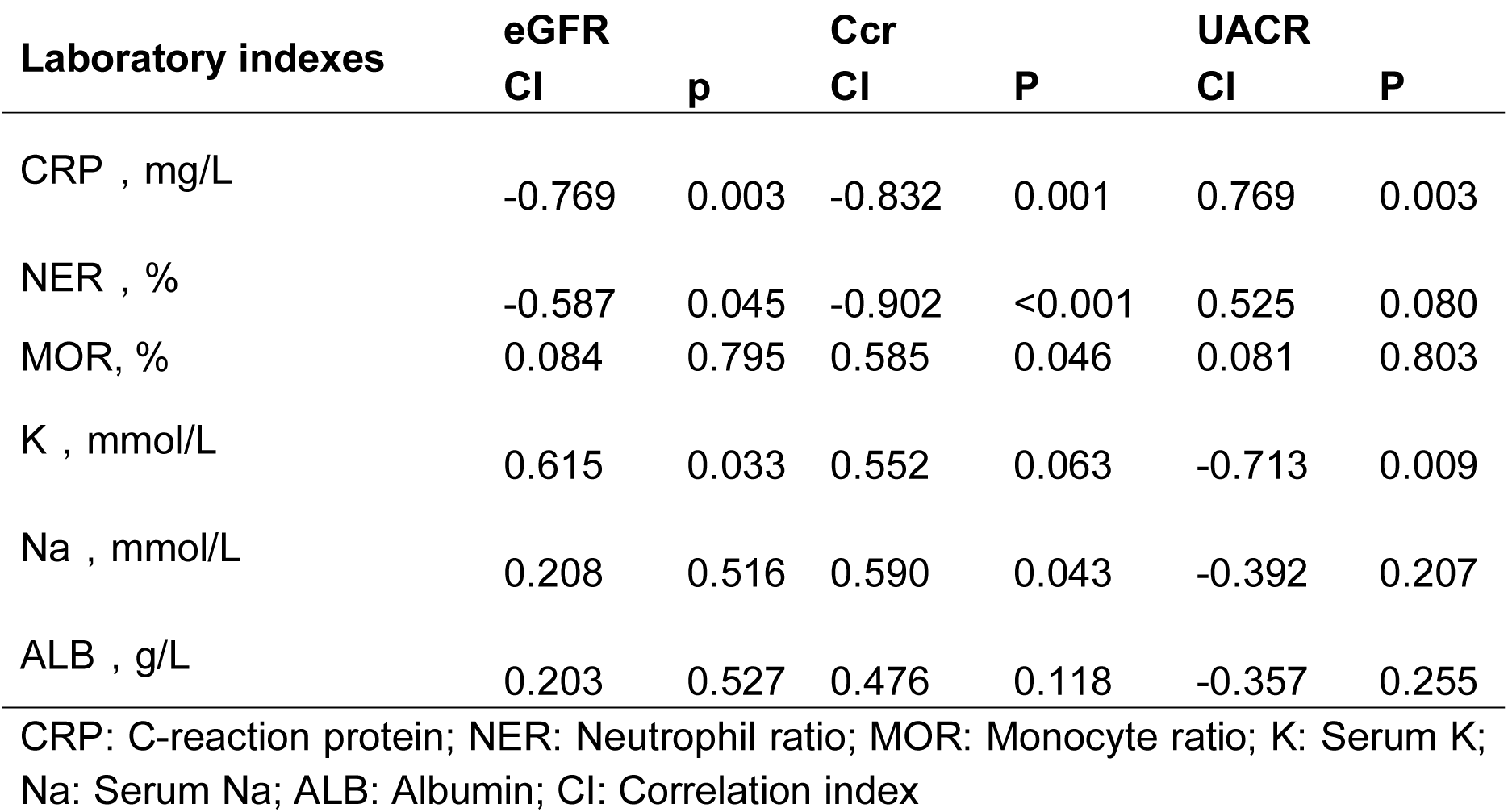
Laboratory indexes related to early renal injury in COVID-19

| **Laboratory indexes** | **eGFR**<br>**CI** | **p** | **Ccr**<br>**CI** | **P** | **UACR**<br>**CI** | **P** |
| --- | --- | --- | --- | --- | --- | --- |
| CRP , mg/L | -0.769 | 0.003 | -0.832 | 0.001 | 0.769 | 0.003 |
| NER , % | -0.587 | 0.045 | -0.902 | <0.001 | 0.525 | 0.080 |
| MOR, % | 0.084 | 0.795 | 0.585 | 0.046 | 0.081 | 0.803 |
| K , mmol/L | 0.615 | 0.033 | 0.552 | 0.063 | -0.713 | 0.009 |
| Na , mmol/L | 0.208 | 0.516 | 0.590 | 0.043 | -0.392 | 0.207 |
| ALB , g/L | 0.203 | 0.527 | 0.476 | 0.118 | -0.357 | 0.255 |

CRP: C-reaction protein; NER: Neutrophil ratio; MOR: Monocyte ratio; K: Serum K; Na: Serum Na; ALB: Albumin; CI: Correlation index

**Figure 2.**
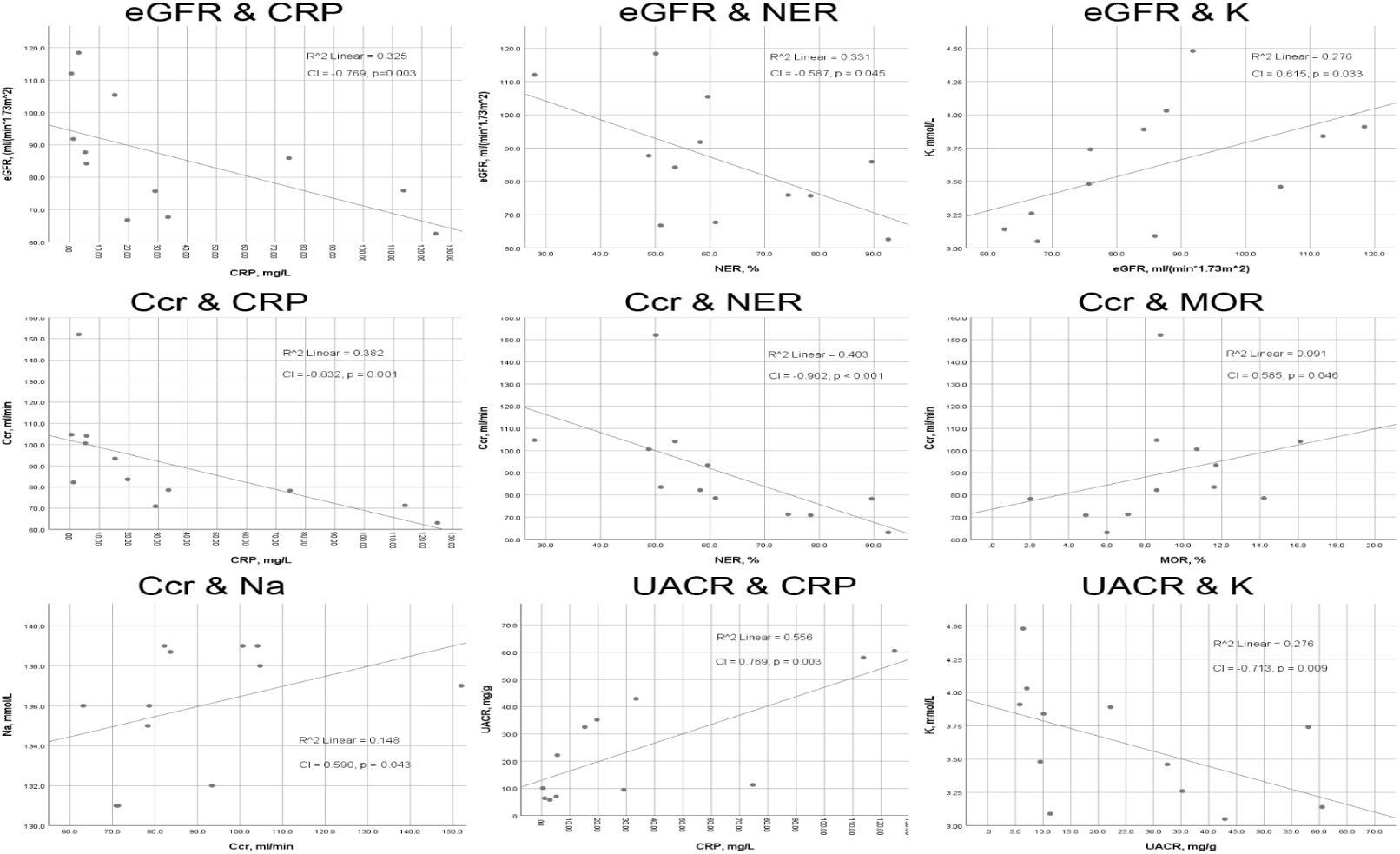
Significant linear correlation between renal function indicators (eGFR, Ccr, UACR) and some biochemical indicators (CRP, NER, MOR, K, Na). (CI: Correlation index; eGFR: estimated glomerular filtration rate; Ccr: endogenous creatinine clearance; UACR: urine microalbumin/creatinine ratio; CRP: C-reactive protein; NER: neutrophil ratio; MOR: monocyte ratio; K: serum potassium; Na: serum sodium)

## Discussion

Coronaviruses are single-stranded positive-strand RNA viruses. In 2014, the International Virological Classification Committee classified them into four genera: α, β, γ, and δ according to serotype and genomic characteristics. There are six known types of coronaviruses that infect humans: α-genus 229E, NL63; β-genus OC43, HKU1, SARSr-CoV and MERSr-CoV.[10] Previous case reports suggest that the spectrum of diseases caused by coronavirus infection is polarized. For example, 229E and NL63 in the α-genus, and OC43 and HKU1 in the β-genus often cause common colds, which are mainly mild, usually without obvious fever, or only low fever. Usually, it is self-limiting and heals without special treatment. However, the other two coronaviruses in the β-genus, SARSr-CoV and MERSr-CoV, often cause acute respiratory syndrome. They mainly cause severe pneumonia, with obvious clinical symptoms, such as fever, dry cough, acute respiratory symptoms and fatigue. Usually, Obvious changes can be found in lung imaging. They are two types of coronavirus pneumonia that require close observation and intensive medical treatment. The 2019-nCov identified this time is a β-genus coronavirus. The diseases caused by its infection combines the characteristics of the previous six coronaviruses, and the disease spectrum is broad.[7]

Peng Zhou et al[11] reported the identification and characterization of 2019-nCoV. Through full-length genome sequences analysis, they found that the whole genome of 2019-nCoV shared 79.5% sequence identify to SARS-CoV. The pairwise protein sequence analysis of seven conserved non-structural proteins showed that this virus belongs to the species of severe acute respirator syndrome-related coronaviruses (SARSr-CoV). In addition, they also confirmed that 2019-nCov used the same cell entry receptor, Angiotensin converting enzyme II (ACE2), as SARS-CoV. The high degree of similarity in gene sequence and cellular mechanism of 2019-nCoV and SARS-CoV suggests that the risk factors of mortality could also be similar. SARS is an acute respiratory infectious disease with pulmonary parenchyma and/or interstitium as the main site of invasion and multiple organ injury. Previous clinical studies have found that, similar to SARS, the causes of death caused by 2019-nCov are not only lung tissue damage, but also heart, liver, kidney and other organ dysfunction or even failure, which is one of the important causes of aggravation and death.[8,9] In the previous SARS case study[12], acute renal injury was found to be the top risk factor of mortality, even higher than acute respiratory distress. In that case study, all patients who eventually died had a progressive rise of Scr, and the rise of Scr was rapid in those who succumbed early in their illness. Previously, an ongoing case study reported 59 patients infected by 2019-nCoV, including 28 severe cases and 3 death. In that study, 63% of the patients exhibited proteinuria, and 19% and 27% of the patients had an elevated level of Scr and BUN respectively. Computed tomography (CT) scans revealed abnormal renal imaging in all patients. The results suggested that renal injury was common in COVID-19, which may contribute to multiorgan failure and death eventually.[13]

In this study, 12 COVID-19 cases were analyzed. Although all the patients still had normal level of Scr and BUN, the high incidence of early renal injury in COVID-19 was found by calculating eGFR, CCR and UACR. The abnormal rates of eGFR, Ccr, and UACR were 66.7%, 41.7%, and 41.7%, respectively. Combined detection of UMA, A1M, IGU, and TRU could be helpful for the diagnosis of early renal injury in COVID-19. Furthermore, the study also found that the degree of early renal injury was significantly related to C-reactive protein (CRP) and neutrophil ratio (NER), suggesting that the more severe the infection, the more obvious the early renal injury. Hypokalemia and hyponatremia were common in patients with COVID-19, and there was a correlation with the degree of renal injury. Xin Zou et al[14] analyzed the single-cell RNA sequencing datasets to explore the expression of ACE2 in the main physiological systems of the human body, including the respiratory, cardiovascular, digestive and urinary system. The study showed that heart, esophagus, kidney, bladder, and ileum have similar or higher ACE2 expression than in alveoli. In the analysis of specific cell types, the expression of ACE2 in renal proximal tubule cells was about four times higher than that of type II alveolar cells (AT2). The results suggest that the kidney may be one of the primary targets of attack for the 2019-nCov.

Scr and BUN are commonly used indicators for the detection of renal function. However, the sensitivity of both of them is poor. Usually, abnormalities occur only when renal function is significantly damaged, as results, they cannot be used as indicators for evaluating early renal injury. Urine microprotein refers to some proteins that are difficult to detect by conventional qualitative or quantitative methods. The detection of urine microprotein can detect the kidney and some other organs’ functional status early. Urine microalbumin is a sensitive indicator of early glomerular injury. Microalbumin has a small molecular weight and is negatively charged. Normally, due to the charge selection barrier of the glomerular filtration membrane, the majority of urine microalbumin cannot pass through the filtration membrane under the action of electrostatic homogeneous repulsion. In the early stage of renal function injury, the charge selectivity of the glomerular filtration membrane decreases, the pore size on the glomerular filtration membrane increases, and the negatively charged structural components of the glomerular filtration membrane change, leading to microalbumin increased excretion from urine.[15] Urine α1-microglobulin is an important indicator for monitoring renal tubular injury. It is a glycoprotein with a molecular weight of 26,000 to 33,000 and is widely distributed in human body. α1-microglobulin can pass through the glomerular filtration membrane, and 95% to 99% is reabsorbed in the proximal tubule, and it is not affected by pH. In the early stage of renal injury, the renal tubule reabsorption function reduced, resulting in urine α1-microglobulin excretion increased, and the elevation is consistent with the degree of renal tubular injury.[16] Due to the large molecular weight, immunoglobulin G is difficult to pass through the glomerular filtration membrane. When renal function is impaired, it can lead to increased permeability of the glomerular filtration membrane. The excretion of albumin in urine increases, and as the lesions worsen, the excretion of immunoglobulin G in urine also increases. Therefore, the detection of urine immunoglobulin G can help to judge the degree of damage on the glomerular filtration membrane.[17] The main physiological function of transferrin is to transport iron ions. It is a negatively charged protein, and its isoelectric point is one unit higher than that of albumin. Normally, it cannot pass through the positively charged glomerular filtration membrane. Urine transferrin is one of the indicators of early glomerular injury, which mainly reflects the damage of glomerular filtration membrane charge selection barrier.[18]

## Strengths and limitations

Strengths of this study are as follows: Firstly, this study first investigated the incidence of early renal injury in COVID-19 and revealed the prevalence of early renal injury in COVID-19 patients. Secondly, this study clarified the value of multi-index combined detection for the diagnosis of early renal injury in COVID-19, and provided a basis for early detection and early intervention. The present study has some limitations that must be taken into account when considering its contribution. First and most significantly is the small sample size of this study. COVID-19 is a newly emerging epidemic. The cases found in this city are all imported cases. After strict prevention and control, there are fewer confirmed cases in this city. Therefore, the number of cases included in this research is also small. In addition, considering COVID-19 is a completely new disease, and we do not yet fully know the characteristics of its cases, the type of research used in this study is designed as a descriptive study, which initially explores the basic characteristics of the disease and does not set the control group.

## Conclusions

This study found that early renal injury was common in patients with COVID-19. Combined detection of UMA, A1M, IGU, and TRU could be helpful for the diagnosis 12 of early renal injury in COVID-19. The 12 cases of COVID-19 reported in this study were all healed and discharged after being treated with antivirals, maintaining water and electrolyte balance, enhancing immunity, oxygen therapy, and traditional chinses medicine treatment. In the process of diagnosis and treatment, the assessment of early injury of important organs and comprehensive protective treatment has played a key role. In particular, in the diagnosis of early renal injury, evidence of COVID-19 with renal injury could be found in time by combining multiple indexes detection. Then, by performing comprehensive treatment measures, such as stopping the use of nephrotoxic drugs timely, maintaining water-electrolyte and acid-base balance and performing effective organ support treatment et al, the complications can be reduced effectively and the prognosis can be improved significantly.

## Data Availability

No additional data are available.

## Acknowledgements

We greatly appreciate all the people, particularly the clinicians, nurses and scientists who are heroically battling to eradicate this disaster for days and nights. We also thank all patients involved in this study.

## Contributors

Hong XW, Chi ZP and Liu GY are joint first authors. Zhang YH obtained funding. Hong XW and Qu LZ designed the study. Huang H, Fan JR, Lin XW, Wu LJ, and Zhou YH collected the data. Chi ZP and Liu GY were involved in data verification and analyzed the data. Guo SQ, Chen RL, Zhang QC, Wu SW, Wang LY, Pan ZQ, Lin H are responsible for the diagnosis and treatment of patients. Hong XW drafted the manuscript. Zhang YH and Qu LZ contributed to the interpretation of the results and critical revision of the manuscript for important intellectual content and approved the final version of the manuscript. All authors have read and approved the final manuscript. Zhang YH is the study guarantor.

## Ethical Approval

This study was approved by the institutional research ethics committee of Shantou Central Hospital.

## Reference

1. Wang C, Horby PW, Hayden FG, et al (2020) A novel coronavirus outbreak of global health concern. Lancet 395 (10223):470–473. doi:10.1016/s0140-6736(20)30185-9

2. Zhu N, Zhang D, Wang W, et al (2020) A Novel Coronavirus from Patients with Pneumonia in China, 2019. The New England journal of medicine 382 (8):727–733. doi:10.1056/NEJMoa2001017

3. Munster VJ, Koopmans M, van Doremalen N, et al (2020) A Novel Coronavirus Emerging in China - Key Questions for Impact Assessment. The New England journal of medicine 382 (8):692–694. doi:10.1056/NEJMp2000929

4. Perlman S (2020) Another Decade, Another Coronavirus. The New England journal of medicine 382 (8):760–762. doi:10.1056/NEJMe2001126

5. Heymann DL, Shindo N (2020) COVID-19: what is next for public health? Lancet 395(10223):542–545. doi:10.1016/s0140-6736(20)30374-3

6. Expert Group of China National Health Commission and Administration of Traditional Chinese Medicine (2020) guidelines for diagnosis and treatment of New Coronavirus Pneumonia (Trial Version 5). Chinese Journal of Integrated Traditional and Western Medicine. doi: http://kns.cnki.net/kcms/detail/11.2787.R.20200208.1034.002.html

7. Wang LH (2020) Characteristics and countermeasures of 2019-nCoV infection. Chinese Journal of Experimental and Clinical Infectious Diseases (Electronic Edition) 14(1):1–5. doi: http://kns.cnki.net/kcms/detail/11.9284.r.20200212.1113.002.html

8. Huang C, Wang Y, Li X, et al (2020) Clinical features of patients infected with 2019 novel coronavirus in Wuhan, China. Lancet 395 (10223):497–506. doi:10.1016/s0140-6736(20)30183-5

9. Chen N, Zhou M, Dong X, et al (2020) Epidemiological and clinical characteristics of 99 cases of 2019 novel coronavirus pneumonia in Wuhan, China: a descriptive study. Lancet 395 (10223):507–513. doi:10.1016/s0140-6736(20)30211-7

10. Zumla A, Chan JF, Azhar EI, et al (2016) Coronaviruses - drug discovery and therapeutic options. Nature reviews Drug discovery 15 (5):327–347. doi:10.1038/nrd.2015.37

11. Zhou P, Yang XL, Wang XG, et al (2020) A pneumonia outbreak associated with a new coronavirus of probable bat origin. Nature. doi:10.1038/s41586-020-2012-7

12. Chu KH, Tsang WK, Tang CS, et al (2005) Acute renal impairment in coronavirus-associated severe acute respiratory syndrome. Kidney international 67 (2):698–705. doi:10.1111/j.1523-1755.2005.67130.x

13. Li Z, Wu M, Guo J, et al (2020) Caution on Kidney Dysfunctions of 2019-nCoV Patients. MedRxiv. doi: https://doi.org/10.1101/2020.02.08.20021212

14. Zou X, Chen K, Zou JW, et al (2020) The single-cell RNA-seq data analysis on the receptor ACE2 expression reveals the potential risk of different human organs vulnerable to Wuhan 2019-nCoV infection. Frontiers Journals of Medicine. doi: https://doi.org/10.1007/s11684-020-0754-0

15. Chen L, Li JX, Huang XB, et al (2011) Determination and its significance of the ratio of urine microalbumin to urine cretinine in patients with nephrolithiasis complicated with renal insufficiency. Journal of Peking University(Health Sciences) 43 (5):757–760. doi: http://xuebao.bjmu.edu.cn/EN/Y2011/V43/I5/757

16. Nordberg J, Allhorn M, Winqvist I, et al (2007) Quantitative and qualitative evaluation of plasma and urine alpha1-microglobulin in healthy donors and patients with different haemolytic disorders and haemochromatosis. Clinica chimica acta 386 (1-2):31–37. doi:10.1016/j.cca.2007.07.017

17. Hou J, Cheng Y, Hou Y, et al (2019) Lower Serum and Higher Urine Immunoglobulin G Are Associated with an Increased Severity of Idiopathic Membranous Nephropathy. Annals of clinical and laboratory science 49 (6):777–784

18. Casanova AG, Vicente-Vicente L, Hernandez-Sanchez MT, et al (2020) Urinary transferrin pre-emptively identifies the risk of renal damage posed by subclinical tubular alterations. Biomedicine & pharmacotherapy 121:109684. doi:10.1016/j.biopha.2019.1096

